# Tracking population mental health before and across stages of the COVID-19 pandemic in young adults

**DOI:** 10.1101/2022.03.24.22272899

**Authors:** Alex S. F. Kwong, Kate Northstone, Rebecca M. Pearson, Andrew M. McIntosh, Nicholas J. Timpson

## Abstract

The SARS-CoV-2 (COVID-19) pandemic has been associated with worsening mental health. Longitudinal studies have monitored changes in mental health from pre-pandemic levels, identifying critical points for mental health as COVID-19 restrictions evolve. Here we highlight changes in depression and anxiety in the UK from pre-pandemic across four pandemic occasions: April and June 2020, January, and July 2021 – corresponding to changes in COVID-19 restrictions. Data were from >5,000 27–29-year-olds from the Avon Longitudinal Study of Parents and Children (ALSPAC). We found that anxiety almost doubled throughout the pandemic compared to pre-pandemic levels and remained high until July 2021 when COVID-19 restrictions were fully lifted. Depression was lower than pre-pandemic levels in April 2020 but increased as the pandemic evolved until July 2021. Women, those with existing mental/physical health conditions and those with economic hardship were most at risk of sustained poorer mental health across the pandemic. Our results highlight the importance of longitudinal studies for tracking mental health during the COVID-19 pandemic and across virus suppression policy changes.

## Introduction

The SARS-CoV-2 (COVID-19) pandemic and related mitigation measures such as lockdown, social distancing and self-isolation have been associated with poor mental health in the early stages of the pandemic, [1] and as it progressed throughout 2020 and 2021. [2] Longitudinal studies have been vital in monitoring mental health across populations and in response to evolving COVID-19 restrictions, with the added benefit of being able to compare pre-pandemic prevalence with that observed throughout the pandemic. [3, 4] This has aided our ability to examine how mental health has changed over time and who may be at greater risk of poorer mental health at key stages throughout the pandemic (i.e., during periods of heightened or relaxed restrictions).

Findings from several longitudinal studies suggest that mental health has deteriorated throughout the pandemic, [5-7] with persistent effects remaining even as restrictions are lifted. [8] This suggests that poorer mental health response to COVID-19 may not be a simple transient reaction to an unprecedented event, but evidence for long-term effects on mental health that may be sustained. [9] However, how severe these effects will be, and the duration is still unclear. Furthermore, changes in mental health are not equally distributed, with women, young people, those with poorer pre-pandemic mental and physical health and those with poorer socioeconomic circumstances purported to be at greater risk during the COVID-19 pandemic. [10] As the response to the pandemic evolves, it is vital that longitudinal studies continue to capture changes in mental health and identify at risk populations to (1) ensure that timely support is provided for the right people, (2) changes in mental health that are coincident with changing restrictions are identified and (3) to identify robust evidence for use in future pandemics.

We present results from a large representative population study of young adults with mental health data before and during the pandemic on four occasions – corresponding to periods of strict and eased COVID-19 restrictions.

## Methods

### Sample

Data were from the Avon Longitudinal Study of Parents and Children (ALSPAC) an ongoing longitudinal population-based cohort study that recruited pregnant women residing in Avon (South-West of England) with expected delivery dates between 1^st^ April 1991 and 31^st^ December 1992. [11, 12] The cohort consists of 14,901 children, now young adults (ages 27-29). [13] Ethical approval for the study was obtained from the ALSPAC Ethics and Law Committee and the Local Research Ethics Committees. This study uses data from four COVID-19 questionnaires assessed between April 2020 (during the first UK national lockdown), June 2020 (when restrictions were partially eased), January 2021 (during the third UK national lockdown) and July 2021 (when UK restrictions were fully eased at this period). [14]

### Mental health measures

Depressive and anxiety symptoms were assessed pre-pandemic and in each COVID-19 questionnaire, examining symptoms within the previous two weeks. Depressive and anxiety symptoms were measured using the Short Mood and Feelings Questionnaire (SMFQ), [15] and Generalised Anxiety Disorder Assessment (GAD-7), [16], respectively. We used established cut-offs to examine the proportion of individuals with probable depression and anxiety (see Supplement). Pre-pandemic depressive symptoms were last measured between 2017-2018 (mean age: ∼25 years old) and anxiety symptoms last measured between 2013-2014 (mean age: ∼22 years old. Further details regarding these measures and additional time points are given in the Supplement. Herein we refer to depressive symptoms as depression and anxiety symptoms as anxiety.

### Statistical analysis

First, we described the prevalence of both depression and anxiety, at the most recent pre-pandemic occasion and across the four COVID-19 data sweeps. We then compared depression and anxiety prevalence across COVID-19 sweeps according to several demographics including sex, pre-pandemic mental and physical health conditions, living alone status, pre-pandemic financial problems, income loss and changes to employment (including furlough). Further information is provided in the Supplement. McNemar’s tests were used to determine differences in prevalence across time and between groups.

## Results

In total, 5,036 participants completed at least one COVID-19 questionnaire, with 3,429 participants completing at least one of the COVID-19 questionnaires *and* the most recent pre-pandemic mental health questionnaire. Sample characteristics of those who completed any COVID-19 questionnaire, or those who completed any COVID-19 questionnaire *and* the latest pre-pandemic assessment, were similar to the most recent pre-pandemic sample in terms of sex, parental education status, deprivation, and pre-pandemic mental health (see Supplement Table 1).

There was evidence that depression decreased from pre-pandemic levels of 23.9% (95% CIs: 22.5-25.4%) to 17.7% (16.2%-19.4%) during the first UK lockdown. However, depression increased to 21.6% (20.1%-23.1%) in the third UK lockdown in December 2020, before decreasing again to 16.1% (14.8%-17.5%) when restrictions were fully lifted in the summer of 2021. Conversely, anxiety increased from pre-pandemic levels of 12.6% (11.4%-14%) to 22.1% (20.3%-24.1%) in the first UK lockdown and rising further to 25% (23.3%-26.8%) during the third UK lockdown, before decreasing to 20.5% (18.8%-22.2%) during the summer of 2021 (see Figure 1; Supplement Tables 2-3).

**Figure 1.**
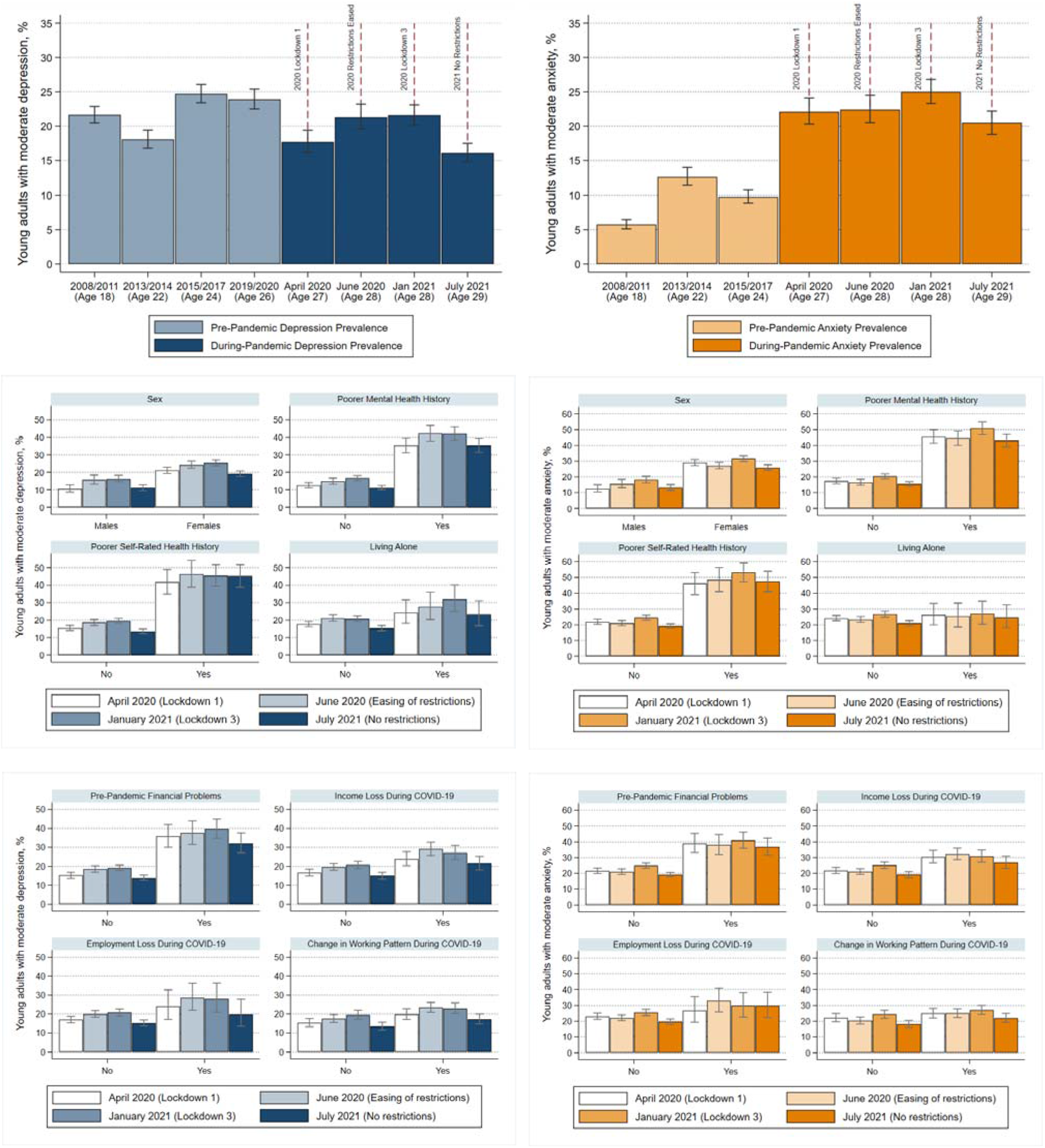
Changes in depression (top left) and anxiety (top right) from before to during COVID-19. Sociodemographic variables by depression (middle left and bottom left) and anxiety (middle right and bottom right) across four waves of COVID-19 data collection. Changes in depression/anxiety from before to during COVID-19 (top panels) were assessed using the sample with both the most recent mental health assessment and any COVID-19 assessment. Changes in depression/anxiety (middle and bottom panels) were assessed using the sample with any COVID-19 measures.

Of those completing at least one COVID-19 questionnaire, women, those with a history of mental/physical health conditions and those with existing financial problems were at greater risk of sustained poorer mental health across the pandemic, compared to their non-at-risk counterparts (Figure). Additionally, unlike their counterparts, those with existing mental/physical health conditions, those with existing financial problems, and those experiencing income/employment loss as result of COVID-19 did not see improvements in anxiety when restrictions were fully eased. A similar pattern was observed such that those with existing physical health conditions and living alone did not see improvements in depression. Full estimates are given in Supplement Tables 4-5. Tests for differences between waves are given in Supplement Tables 6-7. Finally, 30.6% of individuals (95% CIs: 29.4%-31.9%) reported at least one depressive episode (above threshold levels), whilst 37.0% (95% CIs: 35.7%-38.3%) reported at least one anxiety episode across any of the COVID-19 questionnaires (Supplement Tables 8-9).

## Discussion

Using data from a longitudinal population study with pre-pandemic information, we found that mental health changes during COVID-19 in young people are closely aligned to COVID-19 mitigation measures, but that mental health improved once restrictions were fully eased. Whilst mitigation measures are crucial for virus suppression, provisions should be made to ensure that support is available for those with poorer mental health during current and future pandemics.

Our results provide evidence for fluctuations early on in the pandemic, followed by a sustained deterioration of mental health that was not just a transient response to an unprecedented event, These results oppose previous research using more intensively collected data, [1] and show sustained periods of poorer mental health lasting for almost a year, with only clear evidence of improvement during a period of maintained COVID-19 eased restrictions (i.e. summer of 2021).

As shown in previous work, the effects of the COVID-19 pandemic have not been universal, with women, those with existing mental/physical health conditions and those with financial/economic hardship most at risk of poorer mental health. [5, 6, 10] Our results build upon such work highlighting that whilst some populations saw improvements when restrictions were eased, others did not (e.g., those with existing health conditions and those with economic hardship). However, it should be noted that effects differed by depression and anxiety, posing a further complexity for treatment and support by mental health professionals.

Work here has been allowed given the availability of repeated measures of depression and anxiety both pre-and-during the pandemic. We have been able to examine changes in mental health that were coincident with COVID-19 policy changes. However, our study has the potential for bias through missing data and study attrition like any longitudinal study and is weakened by varying follow up times between pre-pandemic and COVID-19 assessments, despite measures taken to address this using a wealth of historical data. Furthermore, our results are specific to young adults and thus may not be generalisable to all populations. However, young adults were posited to be a key group at risk of COVID-19, [17] so detailed analysis of this population is essential as this population is likely to face the social and economic fallout from the pandemic.

The use of longitudinal studies is key for policy makers and health officials in determining how mental has changed from before and during the pandemic, and the identification of who is most at risk and when.

## Data Availability

The ALSPAC study website contains details of all data available through a fully searchable data dictionary (http://www.bristol.ac.uk/alspac/researchers/our-data/).

## Contributors

ASFK, KN, RMP, AMM and NJT contributed to the conception and design of the study. ASFK carried out the study and analysed the data. ASFK drafted the initial output. All authors contributed to the interpretation of data. All authors have read and approved the final version of the manuscript.

ASFK will serve as guarantor for the contents of the paper.

## Acknowledgments

We are extremely grateful to all the families who took part in this study, the midwives for their help in recruiting them, and the whole ALSPAC team, which includes interviewers, computer and laboratory technicians, clerical workers, research scientists, volunteers, managers, receptionists and nurses. Study data were collected and managed using REDCap electronic data capture tools hosted at the University of Bristol (https://projectredcap.org/resources/citations/). REDCap (Research Electronic Data Capture) is a secure, web-based software platform designed to support data capture for research studies The ALSPAC study website contains details of all data available through a fully searchable data dictionary (http://www.bristol.ac.uk/alspac/researchers/our-data/).

