## Supplement for "Tracking population mental health before and across stages of the COVID-19 pandemic in young adults"

**Methods**

**Additional sample information**

Data were from the Avon Longitudinal Study of Parents and Children (ALSPAC) an ongoing longitudinal population-based study that recruited pregnant women residing in Avon (South-West of England) with expected delivery dates between 1st April 1991 and 31st December 1992. [1, 2] The cohort consists of 13,761 mothers and their partners, and their 14,901 children, now young adults. [3] The study website contains details of all data available through a fully searchable data dictionary (http://www.bristol.ac.uk/alspac/researchers/our-data/). Ethical approval for the study was obtained from the ALSPAC Ethics and Law Committee and the Local Research Ethics Committees. This study uses data from the original ALSPAC children's cohort (young adults aged between 27 and 29) who completed four questionnaires about the impact and consequences of the COVID-19 pandemic in April 2020 (during the first UK national lockdown), June 2020 (when restrictions were partially eased), January 2021 (during the third UK national lockdown) and July 2021 (when UK restrictions were fully eased). [4-6]

**Additional mental health measures information**

Depressive symptoms were measured using the Short Mood and Feelings Questionnaire (SMFQ), [7] a 13-item instrument with scores ranging between 0-26 with higher scores indicting higher depressive symptoms. Anxiety symptoms were measured using the Generalised Anxiety Disorder Assessment (GAD-7), [8] a 7-item instrument with scores ranging between 0-21 with higher scores indicting greater generalised anxiety disorder symptoms. Both the SMFQ and GAD-7 have recommended cut-offs for examining the proportion of individuals with probable depression (≥11 on SMFQ; [7]) and generalised anxiety disorder (≥10 on GAD-7; [9]) with good sensitivity and specificity for identifying clinical disorder using validated interviews and instruments. Given the age range for anxiety, we supplemented these results with a more recent measure of probable anxiety assessed by the Clinical Interview Schedule – Revised (CISR), [10] between 2015-2017 (mean age: ~24 years old).

**Demographic information**

For both ALSPAC cohorts, the sex of the participant was recorded at the time of the COVID-19 questionnaire. Where this was missing, historical data from previous questionnaires and clinics was used to identify the sex of the participant. Sex was coded as 0 for males or 1 for females. A poorer history of mental health was assessed using the CIS-R to derive a history of major depressive disorder (MDD) or generalised anxiety disorder (GAD) between 2008-2010 (mean age: ~18 years old) and 2015-2017 (mean age ~24 years old). Responses were coded as 0 for no for both disorders on both occasions and 1 for yes for any disorder on either occasion. Poorer physical health was assessed between 2018-2019 (mean age ~26 years old) by asking participants to rate their health from ‘excellent’, ‘very good’, ‘good’, ‘fair’ and ‘poor’, with poorer physical health coded as 1 when rated ‘fair’ or ‘poor’. Living alone status was assessed at the beginning of the pandemic with responses ranging from 1 as living alone or 0 if participants lived with at least one other person during the first COVID-19 questionnaire. A recent history of financial problems was assessed between 2018-2019 (mean age ~26 years old) and coded as 0 if the participant did not have any recent financial problems in the last 12 months and 1 if they did. Income loss during the COVID-19 pandemic was assessed at the second COVID-19 questionnaire in June 2020 and was coded as 0 if the participant reported being better off or the same financially compared to before the pandemic and 1 if they reported being worse off now compared to before the pandemic. Employment loss was assessed at the second COVID-19 questionnaire in June 2020 and calculated by first asking participants if their employment situation had changed due to COVID-19. Employment loss was then derived by identifying those who reported now being ‘employed but now unemployed’ or ‘were previously working but were now no longer working’. Changes to working patterns such as being on furlough or working increased/reduced hours were derived by again asking if participants’ employment situation had changed due to COVID-19. We coded individuals as having work change if they reported working ‘more hours than before the pandemic’, ‘less hours than before the pandemic’, ‘were employed but on paid leave or furlough’ or ‘employed but on unpaid leave’. Further information on these COVID-19 specific variables is reported elsewhere. [11]

**Statistical analysis**

Additional pre-pandemic time points for the SMFQ and CIS-R anxiety measures were included in figures to aid interpretation of longitudinal trends but are not formally included in these analyses as this would result in a significant reduction of sample size and introduce potential selection biases. Sampling weights are not available in the ALPSAC study and we chose not to impute missing outcome data as we were primarily interested in raw mental health data that could be affected by the virus suppression measures. We estimated differences in proportions above and below thresholds using McNemar’s tests. Results did not vary by restricting to those with pre-pandemic *and* all COVID-19 data (Supplement Tables 2-3). All analyses were conducted in StataMP, version 17 (StataCorp LLC).

**Results**

| **Supplementary Table 1. Participant demographics across the COVID-19 data sweeps and pre-pandemic data** | | | | | | | | | |
| --- | --- | --- | --- | --- | --- | --- | --- | --- | --- |
|  | **Recent PP (n=4020)** | **Q1 (n=2866)** | **Q2 (n=2602)** | **Q3 (n=4056)** | **Q4 (n=3892)** | **Any CV Q (n=5036)** | **All CV Q (n=1764)** | **Any CV + recent PP (n=3429)** | **All CVs + recent PP (n=1514)** |
| **Age  (in years)** | 25.3 (24-27) | 27.6  (27-29) | 27.8  (27-29) | 28.4  (27-29) | 29.0  (28-30) | - | - | - | - |
| **Sex (female)** | 2681 (66.8) | 2050 (71.6%) | 1840 (70.8%) | 2863 (66.2%) | 2570 (66.2%) | 3050 (65.4%) | 1263 (71.7%) | 2353 (68.7%) | 1093 (72.3%) |
| **A level of higher** | 1715 (47.7%) | 1280 (49.6%) | 1192 (50.6%) | 1682 (46.2%) | 1584 (45.5%) | 2063 (45.7%) | 835 (51.2%) | 1493 (48.4%) | 729 (53.1%) |
| **O level** | 1204 (33.5%) | 869 (33.6%) | 785 (33.3%) | 1265 (34.7%) | 1225 (35.2%) | 1566 (34.7%) | 527 (32.9%) | 1046 (33.9%) | 439 (32.0%) |
| **< O level** | 679 (18.9%) | 434 (16.8%) | 379 (16.1%) | 697 (19.1%) | 673 (19.3%) | 881 (19.5%) | 239 (14.9%) | 549 (17.8%) | 206 (15.0%) |
| **Least Deprived** | 1412 (38.2%) | 1053 (39.6%) | 924 (38.4%) | 1396 (37.1%) | 1330 (37.1%) | 1718 (37.0%) | 650 (39.6%) | 1228 (38.7%) | 568 (40.3%) |
| **Most Deprived** | 245 (6.6%) | 161 (6.1%) | 149 (6.2%) | 261 (6.9%) | 233 (6.5%) | 329 (7.1%) | 92 (5.6%) | 205 (6.5%) | 75 (5.3%) |
| **Any MH** | 642 (19.7%) | 506 (20.7%) | 456 (20.6%) | 623 (19.3%) | 580 (18.8%) | 756 (19.3%) | 311 (20.1%) | 567 (19.6%) | 280 (20.5%) |

PP refers to most recent pre-pandemic assessment of mental health. Educational attainment is based upon parental measures of educational attainment.

| **Supplementary Table 2. Depression prevalence (top) and summary scores (bottom)** | | | | |
| --- | --- | --- | --- | --- |
| **Percentage of Individuals Above Threshold (95% CIs)** | | | | |
| **Recent PP** | **COVID-19 Q1** | **COVID-19 Q2** | **COVID-19 Q3** | **COVID-19 Q4** |
| **Raw** |  |  |  |  |
| 24.4 (23.1-25.7) | 18.1 (16.8-19.6) | 21.8 (20.3-23.5) | 22.3 (21-23.6) | 16.6 (15.4-17.8) |
| n=4020 | n=2812 | n=2567 | n=3978 | n=3794 |
| **Restricted to PP and any COVID data** | |  |  |  |
| 23.9 (22.5-25.4) | 17.7 (16.2-19.4) | 21.3 (19.6-23.2) | 21.6 (20.1-23.1) | 16.1 (14.8-17.5) |
| n=3414 | n=2219 | n=2030 | n=2875 | n=2786 |
| **Restricted to PP and all COVID data** | |  |  |  |
| 24 (21.9-26.3) | 16.9 (15.1-19) | 20.7 (18.8-23) | 19.9 (17.9-22.7) | 14.6 (12.9-16.5) |
| n=1424 | n=1424 | n=1424 | n=1424 | n=1424 |

| **Mean scores (SD)** |  |  |  |  |
| --- | --- | --- | --- | --- |
| **Recent PP** | **COVID-19 Q1** | **COVID-19 Q2** | **COVID-19 Q3** | **COVID-19 Q4** |
| **Raw** |  |  |  |  |
| 6.9 (6.4) | 6.2 (5.6) | 6.6 (5.7) | 6.7 (6.1) | 5.1 (5.8) |
| n=4020 | n=2812 | n=2567 | n=3978 | n=3794 |
| **Restricted to PP and any COVID data** | |  |  |  |
| 6.8 (6.4) | 6.2 (5.6) | 6.5 (5.7) | 6.6 (6) | 5.0 (5.7) |
| n=3414 | n=2219 | n=2030 | n=2875 | n=2786 |
| **Restricted to PP and all COVID data** | |  |  |  |
| 6.9 (6.3) | 6.0 (5.5) | 6.3 (5.6) | 6.3 (5.9) | 5.0 (5.5) |
| n=1424 | n=1424 | n=1424 | n=1424 | n=1424 |

PP refers to most recent pre-pandemic assessment. Raw refers to the threshold/scores for just that occasion in isolation.

| **Supplementary Table 3. Anxiety prevalence (top) and summary scores (bottom)** | | | | |
| --- | --- | --- | --- | --- |
| **Percentage of Individuals Above Threshold (95% CIs)** | | | | |
| **Recent PP** | **COVID-19 Q1** | **COVID-19 Q2** | **COVID-19 Q3** | **COVID-19 Q4** |
| **Raw** |  |  |  |  |
| 12.9 (11.9-14.2) | 24.4 (22.8-26) | 23.8 (22.2-25.5) | 27.1 (25.7-28.5) | 21.7 (20.4-23) |
| n=3339 | n=2850 | n=2571 | n=4010 | n=3835 |
| **Restricted to PP and any COVID data** | |  |  |  |
| 12.6 (11.4-14) | 22.1 (20.3-24.1) | 22.4 (20.5-24.5) | 25 (23.3-26.8) | 20.5 (18.8-22.2) |
| n=2697 | n=1811 | n=1676 | n=2289 | n=2204 |
| **Restricted to PP and all COVID data** | |  |  |  |
| 11.9 (10.2-13.8) | 20.8 (18.6-23.1) | 21.9 (19.7-24.3) | 23.4 (21.1-25.8) | 19.3 (17.2-21.6) |
| n=1224 | n=1224 | n=1224 | n=1224 | n=1224 |

| **Mean scores (SD)** |  |  |  |  |
| --- | --- | --- | --- | --- |
| **Recent PP** | **COVID-19 Q1** | **COVID-19 Q2** | **COVID-19 Q3** | **COVID-19 Q4** |
| **Raw** |  |  |  |  |
| 4.6 (4.6) | 6.2 (5.3) | 6.1 (5.3) | 6.7 (5.5) | 5.7 (5.2) |
| n=3339 | n=2850 | n=2571 | n=4010 | n=3825 |
| **Restricted to PP and any COVID data** | |  |  |  |
| 4.5 (4.5) | 5.9 (5.1) | 5.8 (5.1) | 6.4 (5.3) | 5.5 (5.1) |
| n=2697 | n=1811 | n=1676 | n=2289 | n=2204 |
| **Restricted to PP and all COVID data** | |  |  |  |
| 4.5 (4.4) | 5.8 (5.1) | 5.7 (5) | 6.2 (5.2) | 5.4 (5.0) |
| n=1224 | n=1224 | n=1224 | n=1224 | n=1224 |

PP refers to most recent pre-pandemic assessment. Raw refers to the threshold/scores for just that occasion in isolation.

| **Supplementary Table 4. Prevalence in depression by varying demographics in individuals who completed at least one COVID-19 questionnaire** | | | | |
| --- | --- | --- | --- | --- |
|  | **Percentage Above Threshold (95% CIs)** | | | |
|  | **COVID-19 Q1** | **COVID-19 Q2** | **COVID-19 Q3** | **COVID-19 Q4** |
| **Men** | 10.6 (8.7-13.0) | 15.7 (13.3-18.5) | 16.2 (14.4-18.3) | 11.2 (9.5-13.0) |
|  | n=800 | n=751 | n=1342 | n=1273 |
| **Women** | 21.1 (19.4-23.0) | 24.3 (22.4-26.4) | 25.4 (23.7-27.1) | 19.2 (17.7-20.8) |
|  | n=2009 | n=1813 | n=2633 | n=2510 |
| **No Pre-Existing MH** | 12.6 (11.2-14.2) | 14.9 (13.3-16.7) | 16.6 (15.2-18.1) | 11.1 (9.9-12.5) |
|  | n=1899 | n=1742 | n=2548 | n=2433 |
| **Yes Pre-Existing MH** | 35.4 (31.3-39.7) | 42.3 (37.8-46.9) | 42.1 (38.2-46.1) | 35.3 (31.5-39.4) |
|  | n=500 | n=447 | n=613 | n=566 |
| **No Pre-Existing SRH** | 15.5 (14.1-17.1) | 18.6 (17.0-20.4) | 19.5 (18.1-21.0) | 13.6 (12.4-14.9) |
|  | n=2248 | n=2054 | n=2953 | n=2842 |
| **Yes Pre-Existing SRH** | 41.7 (34.8-48.9) | 46.4 (38.9-54.1) | 45.5 (39.5-51.7) | 45.2 (38.9-51.7) |
|  | n=187 | n=166 | n=257 | n=230 |
| **No Live Alone** | 17.7 (16.3-19.2) | 21.1 (19.4-23.0) | 20.8 (19.2-22.5) | 15.4 (13.9-17.0) |
|  | n=2632 | n=1946 | n=2276 | n=2142 |
| **Live Alone** | 24.2 (18.2-31.5) | 27.6 (20.4-36.0) | 31.9 (24.8-40.1) | 23.2 (16.8-31.0) |
|  | n=161 | n=127 | n=144 | n=138 |
| **No Financial Problems** | 15.2 (13.7-16.8) | 18.5 (16.8-20.3) | 19.1 (17.7-20.6) | 13.9 (12.7-15.3) |
|  | n=2125 | n=1943 | n=2762 | n=2675 |
| **Financial Problems** | 35.8 (30.0-42.0) | 37.6 (31.6-44.0) | 39.8 (34.7-45.0) | 32.1 (27.1-37.6) |
|  | n=246 | n=234 | n=347 | n=308 |
| **No Income Loss** | 16.6 (14.8-18.5) | 19.6 (17.9-21.4) | 20.7 (18.8-22.6) | 15.0 (13.3-16.8) |
|  | n=1555 | n=1913 | n=1714 | n=1613 |
| **Income Loss** | 23.8 (20.3-27.7) | 29.1 (25.7-32.8) | 27.1 (23.5-31.0) | 21.5 (18.1-25.2) |
|  | n=500 | n=625 | n=539 | n=517 |
| **No Employ Loss** | 17.1 (15.4-18.9) | 20.0 (18.4-21.8) | 20.8 (19.0-22.7) | 15.2 (13.7-16.9) |
|  | n=1777 | n=2177 | n=1934 | n=1851 |
| **Employ Loss** | 24.1 (17.2-32.8) | 28.6 (22.0-36.3) | 28.0 (21.0-36.4) | 19.7 (13.5-27.8) |
|  | n=116 | n=154 | n=132 | n=122 |
| **No Work Change** | 15.4 (13.3-17.7) | 17.6 (15.6-19.8) | 19.5 (17.3-22.0) | 13.5 (11.6-15.7) |
|  | n=1035 | n=1266 | n=1131 | n=1086 |
| **Work Change** | 19.9 (17.2-22.8) | 23.5 (20.9-26.2) | 22.9 (20.3-25.9) | 17.3 (14.8-20.0) |
|  | n=795 | n=985 | n=868 | n=823 |

MH refers to pre-existing mental health. SRH refers to pre-existing poorer self-rated health.

| **Supplementary Table 5. Prevalence in anxiety by varying demographics in individuals who completed at least one COVID-19 questionnaire** | | | | |
| --- | --- | --- | --- | --- |
|  | **Percentage Above Threshold (95% CIs)** | | | |
|  | **COVID-19 Q1** | **COVID-19 Q2** | **COVID-19 Q3** | **COVID-19 Q4** |
| **Men** | 12.6 (10.5-15.1) | 15.6 (13.2-18.4) | 18.2 (16.3-20.4) | 13.2 (11.4-15.1) |
|  | n=809 | n=751 | n=1355 | n=1289 |
| **Women** | 29.0 (27.1-31.1) | 27.1 (25.1-29.2) | 31.6 (29.9-33.4) | 25.9 (24.2-27.7) |
|  | n=2038 | n=1817 | n=2652 | n=2535 |
| **No MH** | 17.4 (15.8-19.2) | 16.6 (14.9-18.5) | 20.4 (18.8-22.0) | 15.5 (14.1-17.0 |
|  | n=1930 | n=1733 | n=2575 | n=2456 |
| **Yes MH** | 45.6 (41.3-50.0) | 44.6 (40.1-49.2) | 50.8 (46.9-54.7) | 43.1 (39.1-47.1) |
|  | n=502 | n=455 | n=618 | n=576 |
| **No SRH** | 21.9 (20.3-23.7) | 21.0 (19.3-22.8) | 24.5 (23.0-26.1) | 19.1 (17.7-20.5) |
|  | n=2277 | n=2050 | n=2978 | n=2866 |
| **Yes SRH** | 46.1 (39.1-53.2) | 48.5 (41.0-56.1) | 53.2 (47.2-59.2) | 47.4 (41.0-53.9) |
|  | n=191 | n=167 | n=263 | n=232 |
| **No Live Alone** | 24.2 (22.6-25.8) | 23.3 (21.5-25.2) | 26.6 (24.9-28.5) | 20.9 (19.2-22.7) |
|  | n=2666 | n=1948 | n=2286 | n=2172 |
| **Live Alone** | 26.2 (20.0-33.5) | 25.4 (18.5-33.8) | 26.9 (20.3-34.8) | 24.6 (18.1-32.6) |
|  | n=164 | n=126 | n=145 | n=138 |
| **No Financial Problems** | 21.6 (19.9-23.4) | 21.0 (19.3-22.9) | 24.9 (23.3-26.6) | 19.1 (17.7-20.6) |
|  | n=2151 | n=1943 | n=2790 | n=2697 |
| **Financial Problems** | 39.0 (33.2-45.3) | 38.1 (32.0-44.6) | 40.9 (35.9-46.2) | 36.8 (31.6-42.3) |
|  | n=251 | n=231 | n=347 | n=310 |
| **No Income Loss** | 21.8 (19.9-23.9) | 21.1 (19.4-23.0) | 25.2 (23.2-27.2) | 19.2 (17.3-21.1) |
|  | n=1572 | n=1915 | n=1723 | n=1655 |
| **Income Loss** | 30.4 (26.6-34.6) | 32.3 (28.7-36.0) | 30.8 (27.1-34.8) | 26.9 (23.2-30.9) |
|  | n=506 | n=626 | n=548 | n=521 |
| **No Employ Loss** | 23.1 (21.2-25.1) | 22.1 (20.4-23.9) | 25.5 (23.6-27.5) | 19.7 (17.9-21.5) |
|  | n=1795 | n=2178 | n=1951 | n=1877 |
| **Employ Loss** | 26.7 (19.4-35.6) | 32.9 (25.8-40.8) | 29.9 (22.7-38.2) | 29.8 (22.4-38.5) |
|  | n=116 | n=152 | n=134 | n=124 |
| **No Work Change** | 22.2 (19.7-24.8) | 20.4 (18.2-22.7) | 24.3 (21.9-26.9) | 18.1 (16.0-20.5) |
|  | n=1047 | n=1272 | n=1146 | n=1104 |
| **Work Change** | 24.9 (22.1-28.1) | 25.0 (22.4-27.8) | 27.0 (24.2-30.1) | 22.0 (19.3-24.9) |
|  | n=802 | n=979 | n=870 | n=833 |

MH refers to pre-existing mental health. SRH refers to pre-existing poorer self-rated health.

| **Supplementary Table 6. Differences in depression proportions across COVID-19 waves, by varying demographics** | | | | | | |
| --- | --- | --- | --- | --- | --- | --- |
|  | **Prop diff (95% CIs), *x*^2^, *p*** | | | | | |
|  | **COVID Q1 vs Q2** | **COVID Q1 vs Q3** | **COVID Q1 vs Q4** | **COVID Q2 vs Q3** | **COVID Q2 vs Q4** | **COVID Q3 vs Q4** |
| **Overall** | -0.03 (-0.05, -0.02), 16.3, p=0.0001 | -0.03 (-0.05, -0.01), 13.7, p=0.0003 | 0.01 (-0.006, 0.03), 1.6, p=0.223 | 0.002 (-0.02, 0.02), 0.1, p=0.839 | 0.05 (0.03, 0.07), 31.4, p<0.0001 | 0.05 (0.04,0.06), 48.5, p<0.0001 |
| **Men** | -0.05 (-0.08, -0.02), 10.3, p=0.001 | -0.04 (-0.07, -0.01), 7.0, p=0.010 | -0.01 (-0.04, 0.02), 0.5, p=0.55 | 0.003 (-0.03, 0.03), 0.1, p=0.915 | 0.04 (0.01, 0.06), 9.6, p=0.003 | 0.04 (0.02, 0.07), 16.4, p=0.0001 |
| **Women** | -0.03 (-0.05, -0.01), 8.1, p=0.005 | -0.03 (-0.05, -0.01), 7.7, p=0.007 | 0.02 (-0.002, 0.04), 3.2, p=0.086 | 0.002 (-0.02, 0.02), 0.03, p=0.909 | 0.05 (0.03, 0.07), 22.6, p<0.0001 | 0.05 (0.03, 0.07), 32.7, p<0.0001 |
| **No MH** | -0.03 (-0.05, -0.01), 10.2, p=0.001 | -0.03 (-0.05, -0.01), 9.9, p=0.002 | 0.004 (-0.01, 0.02), 0.2, p=0.726 | -0.007 (-0.03, 0.01), 0.4, p=0.561 | 0.04 (0.02, 0.05), 13.1, p=0.0004 | 0.05 (0.03, 0.7), 26.3, p<0.0001 |
| **Yes MH** | -0.06 (-0.11, -0.01), 5.6, p=0.018 | -0.04 (-0.09, 0.01), 2.4, p=0.149 | 0.02 (-0.03, 0.07), 0.6, p=0.493 | 0.01 (-0.04, 0.06), 0.2, p=0.757 | 0.08 (0.02, 0.13), 7.5, p=0.008 | 0.06 (0.01, 0.11), 5.5, p=0.0232 |
| **No SRH** | -0.03 (-0.05, -0.01), 12.5, p=0.0004 | -0.03 (-0.05, -0.01), 12.7, p=0.0004 | 0.02 (-0.001, 0.03), 3.7, p=0.063 | -0.001 (-0.02, 0.02), 0.01, p=0.955 | 0.05 (0.03, 0.07), 30.6, p<0.0001 | 0.06 (0.04, 0.07), 49.1, p<0.0001 |
| **Yes SRH** | -0.05 (-0.14, 0.05), 1.0, p=0.330 | 0.02 (-0.07, 0.10), 0.2, p=0.766 | -0.03 (-0.12, 0.06), 0.42, p=0.627 | 0.04 (-0.03, 0.12), 1.4, p=0.327 | -0.01 (-0.10, 0.07), 0.1, p=0.855 | 0.01 (-0.06, 0.08), 0.1, p=0.883 |
| **No Live Alone** | -0.03 (-0.05, -0.02), 15.0, p=0.0001 | -0.03 (-0.05, -0.01), 11.1, p=0.001 | 0.01 (-0.005, 0.03), 1.9, p=0.184 | 0.009 (-0.01, 0.03), 0.7, p=0.423 | 0.05 (0.03, 0.07), 26.1, p<0.0001 | 0.05 (0.03, 0.07), 28.7, p<0.0001 |
| **Live Alone** | -0.05 (-0.12, 0.02), 2.3, p=0.210 | -0.07 (-0.15, 0.01), 3.85, p=0.076 | -0.01 (-0.10, 0.07), 0.2, p=0.845 | -0.03 (-0.12, 0.05), 0.8, p=0.503 | 0.03 (-0.05, 0.1), 0.5, p=0.629 | 0.04 (-0.05, 0.12),1.0, p=0.424 |
| **No Financial Problems** | -0.03 (-0.05, -0.02), 13.6, p=0.0002 | -0.03 (-0.05, -0.01), 10.65, p=0.001 | 0.01 (-0.006, 0.03), 1.8, p=0.198 | 0.003 (-0.02, 0.02), 0.1, p=0.765 | 0.05 (0.03, 0.07), 26.1, p<0.0001 | 0.05 (0.03, 0.07), 38.2, p<0.0001 |
| **Financial Problems** | -0.02 (-0.09, 0.06), 0.2, p=0.755 | -0.01 (-0.09, 0.06), 0.15, p=0.795 | 0.03 (-0.04, 0.11), 0.8, p=0.451 | -0.01 (-0.08, 0.06), 0.1, p=0.883 | 0.03 (-0.04, 0.1), 1.0, p=0.405 | 0.06 (-0.01, 0.12), 3.9, p=0.064 |
| **No Income Loss** | -0.03 (-0.05, -0.01), 10.3, p=0.002 | -0.03 (-0.06, -0.01), 10.4, p=0.002 | 0.01 (-0.01, 0.03), 1.0, p=0.365 | -0.004 (-0.02, 0.02), 0.1, p=0.768 | 0.05 (0.03, 0.07), 21.7, p<0.0001 | 0.04 (0.02, 0.06), 18.6, p<0.0001 |
| **Income Loss** | -0.05 (0.09, -0.01), 6.26, p=0.016 | -0.02 (-0.06, 0.02), 0.95, p=0.386 | 0.02 (-0.02, 0.06), 0.7, p=0.470 | 0.02 (-0.02, 0.06), 0.8, p=0.426 | 0.06 (0.02, 0.1), 9.9, p=0.002 | 0.05 (0.02, 0.10), 9.7, p=0.0024 |
| **No Employ Loss** | -0.03 (-0.05, -0.01), 12.7, p=0.0004 | -0.03 (-0.05, -0.01), 10.8, p=0.001 | 0.007 (-0.01, 0.03), 0.5, p=0.507 | -0.005 (-0.02, 0.01), 0.3, p=0.617 | 0.04 (0.03, 0.06), 21.9, p<0.0001 | 0.04 (0.03, 0.06), 22.1, p<0.0001 |
| **Employ Loss** | -0.02 (-0.10, 0.07), 0.2, p=0.824 | 0.01 (-0.08, 0.10), 0.1, p=1.0 | 0.08 (-0.01, 0.2), 3.6, p=0.096 | 0 (-0.09, 0.09), 0, p=1.0 | 0.07 (-0.01, 0.16), 3.5, p=0.093 | 0.10 (0.01, 0.19), 3.2, p=0.0266 |
| **No Work Change** | -0.02 (-0.05, 0.001), 3.92, p=0.058 | -0.04 (-0.06, -0.01), 7.0, p=0.009 | 0.01 (-0.01, 0.04), 0.8, p=0.415 | -0.02 (-0.04, 0.01), 1.6, p=0.237 | 0.04 (0.01, 0.06), 10.0, p=0.002 | 0.05 (0.02, 0.08), 15.8, p=0.0001 |
| **Work Change**  MH refers to pre-existing mental health. SRH refers to pre-existing poorer self-rated health. | -0.04 (-0.07, -0.01), 8.3, p=0.004 | -0.03 (-0.06, 0.004), 3.2, p=0.09 | 0.009 (-0.02, 0.04), 0.3, p=0.637 | 0.008 (-0.02, 0.04), 0.3, p=0.634 | 0.06 (0.03, 0.08), 14.8, p=0.0002 | 0.05 (0.02, 0.08), 10.7, p=0.0014 |

| **Supplementary Table 7. Differences in anxiety proportions across COVID-19 waves, by varying demographics** | | | | | | |
| --- | --- | --- | --- | --- | --- | --- |
|  | **Prop diff (95% CIs), *x*^2^, *p*** | | | | | |
|  | **COVID Q1 vs Q2** | **COVID Q1 vs Q3** | **COVID Q1 vs Q4** | **COVID Q2 vs Q3** | **COVID Q2 vs Q4** | **COVID Q3 vs Q4** |
| **Overall** | 0.008 (-0.01, 0.03), 0.7, p=0.420 | -0.02 (-0.04, -0.004), 6.2, p=0.014 | 0.02 (0.004, 0.04), 5.9, p=0.017 | -0.03 (-0.05, -0.01), 10.2, p=0.002 | 0.02 (0.006, 0.04), 7.2, p=0.009 | 0.05 (0.03, 0.06), 34.3, p<0.0001 |
| **Men** | -0.03 (-0.06, -0.005), 5.7, p=0.023 | -0.05 (-0.08, -0.02), 13.2, p=0.0004 | -0.01 (-0.04, 0.01), 1.0, p=0.368 | -0.02 (-0.05, 0.007), 2.6, p=0.129 | 0.03 (-0.002, 0.06), 3.7, p=0.071 | 0.05 (0.02, 0.07), 16.9, p<0.0001 |
| **Women** | 0.02 (0.002, 0.05), 5.0, p=0.029 | -0.01 (-0.03, 0.01), 0.9, p=0.360 | 0.04 (0.01, 0.06), 10.2, p=0.002 | -0.03 (-0.06, -0.009), 7.9, p=0.006 | 0.02 (0.003, 0.05), 4.2, p=0.047 | 0.05 (0.03, 0.07), 20.6, p<0.0001 |
| **No MH** | 0.008 (-0.01, 0.03), 0.7, p=0.434 | -0.03 (-0.05, -0.009), 8.31, p=0.005 | 0.01 (-0.009, 0.03), 1.2, p=0.299 | -0.03 (-0.06, -0.01), 9.9, p=0.002 | 0.01 (-0.01, 0.03), 1.2, p=0.303 | 0.05 (0.03, 0.06), 25.1, p<0.0001 |
| **Yes MH** | -0.01 (-0.07, 0.03), 0.3, p=0.675 | -0.03 (-0.08, 0.02), 1.6, p=0.235 | 0.03 (-0.03, 0.08), 1.0, p=0.371 | -0.03 (-0.08, 0.02), 1.5, p=0.266 | 0.04 (-0.01, 0.10), 2.9, p=0.111 | 0.05 (-0.01, 0.10), 3.4, p=0.079 |
| **No SRH** | 0.006 (-0.01, 0.03), 0.4, p=0.547 | -0.02 (-0.04, -0.0006), 4.3, p=0.044 | 0.03 (0.005, 0.05), 6.6, p=0.012 | -0.03 (-0.05, -0.009), 8.3, p=0.005 | 0.03 (0.01, 0.05), 8.1, p=0.005 | 0.05 (0.03, 0.07), 31.8, p<0.0001 |
| **Yes SRH** | 0 (-0.09, 0.09), 0, p=1.0 | -0.04 (-0.12, 0.04), 1.1, p=0.360 | 0.01 (-0.08, 0.11), 0.1, p=0.880 | -0.03 (-0.11, 0.06), 0.5, p=0.608 | 0.007 (-0.09, 0.10), 0.03, p=1.0 | 0.06 (-0.02, 0.14), 2.5, p=0.148 |
| **No Live Alone** | 0.006 (-0.01, 0.03), 0.4, p=0.545 | -0.02 (-0.04, -0.004), 5.9, p=0.017 | 0.02 (0.004, 0.04), 6.1, p=0.02 | -0.03 (-0.05, -0.006), 6.7, p=0.011 | 0.03 (0.005, 0.05), 6.1, p=0.016 | 0.05 (0.03, 0.07), 23.5, p<0.0001 |
| **Live Alone** | 0.02 (-0.04, 0.09), 0.6, p=0.607 | -0.007 (-0.08, 0.06), 0.1, p=1.0 | 0 (-0.08, 0.08), 0, p=1.0 | -0.03 (-0.10, 0.05), 0.6, p=0.607 | -0.009, (-0.09, 0.08), 0.1, p=1.0 | 0.01 (-0.08, 0.09), 0.04, p=0.842 |
| **No Financial Prob** | 0.002 (-0.02, 0.02), 0.1, p=0.848 | -0.02 (-0.04, -0.003), 5.6, p=0.021 | 0.03 (0.005, 0.05), 6.3, p=0.014 | -0.03 (-0.05, -0.007), 7.2, p=0.009 | 0.03 (0.007, 0.05), 7.3, p=0.008 | 0.05 (0.03, 0.07), 33.5, p<0.0001 |
| **Financial Problems** | 0.03 (-0.05, 0.11), 0.5, p=0.576 | -0.005 (-0.08, 0.07), 0.02, p=1.0 | 0.03 (-0.05, 0.10), 0.6, p=0.552 | -0.05 (-0.12, 0.02), 1.9, p=0.212 | 0.02 (-0.06, 0.09), 0.2, p=0.749 | 0.03 (-0.03, 0.10), 1.1, p=0.356 |
| **No Income Loss** | 0.01 (-0.006, 0.04), 2.2, p=0.159 | -0.03 (-0.05, -0.009), 7.9, p=0.006 | 0.02 (-0.002, 0.04), 3.4, p=0.077 | -0.04 (-0.06, -0.02), 14.3, p=0.0002 | 0.02 (-0.003, 0.04), 3.1, p=0.091 | 0.05 (0.03, 0.07), 19.0, p<0.0001 |
| **Income Loss** | -0.02 (-0.06, 0.02), 1.1, p=0.353 | 0.009 (-0.04, 0.05), 0.2, p=0.764 | 0.02 (-0.03, 0.07), 0.9, p=0.387 | -0.002 (-0.04, 0.04), 0.01, p=1.0 | 0.04 (0.002, 0.09), 4.7, p=0.038 | 0.04 (-0.01, 0.09), 3.0, p=0.099 |
| **No Employ Loss** | 0.02 (-0.003, 0.04), 2.9, p=0.101 | -0.03 (-0.05, -0.004), 5.8, p=0.018 | 0.03 (0.005, 0.05), 6.1, p=0.016 | -0.04 (-0.06, -0.02), 13.4, p=0.0003 | 0.02 (0.002, 0.04), 4.8, p=0.033 | 0.05 (0.03, 0.07), 23.4, p<0.0001 |
| **Employ Loss** | -0.08 (-0.16, -0.0001), 4.8, p=0.049 | 0.01 (-0.09, 0.11), 0.1, p=1.0 | -0.06 (-0.17, 0.05), 1.4, p=0.327 | 0.02 (-0.07, 0.12), 0.3, p=0.736 | 0.02 (-0.07, 0.11), 0.2, p=0.845 | 0.0 (-0.11, 0.11), 0, p=1.0 |
| **No Work Change** | 0.02 (-0.006, 0.04), 2.4, p=0.140 | -0.03 (-0.06, 0.0008), 3.9, p=0.057 | 0.03 (-0.0002, 0.06), 4.1, p=0.052 | -0.04 (-0.06, -0.01), 8.6, p=0.004 | 0.02 (-0.008, 0.04), 1.9, p=0.185 | 0.05 (0.03, 0.08), 15.2, p=0.0001 |
| **Work Change** | 0.01 (-0.02, 0.04), 0.5, p=0.546 | -0.01 (-0.05, 0.02), 0.7, p=0.458 | 0.03 (-0.009, 0.06), 2.4, p=0.148 | -0.03 (-0.06, 0.005), 3.0, p=0.099 | 0.03 (-0.003, 0.06), 3.4, p=0.077 | 0.05 (0.01, 0.8), 7.7, p=0.0068 |

MH refers to pre-existing mental health. SRH refers to pre-existing poorer self-rated health.

| **Supplementary Table 8. Number of depression occasions (above threshold) across all four COVID-19 assessments** | | | | | | |
| --- | --- | --- | --- | --- | --- | --- |
| **Percentage of Individuals (95% CIs)** | | | | | | |
|  | **Any depression occasion** | **0 occasions** | **1 occasion** | **2 occasions** | **3 occasions** | **4 occasions** |
| **Overall** | 30.6 (29.5, 31.9) | 69.4 (68.1, 70.1) | 17.2 (16.2, 18.3) | 7.7 (7.0, 8.5) | 3.7 (3.2, 4.3) | 2.0 (1.6, 2.4) |
|  | n=1532 | n=3470 | n=862 | n=385 | n=187 | n=98 |

| **Supplementary Table 9. Number of anxiety occasions (above threshold) across all four COVID-19 assessments** | | | | | | |
| --- | --- | --- | --- | --- | --- | --- |
| **Percentage of Individuals (95% CIs)** | | | | | | |
|  | **Any anxiety occasion** | **0 occasions** | **1 occasion** | **2 occasions** | **3 occasions** | **4 occasions** |
| **Overall** | 37.0 (35.7, 38.3) | 63.0 (61.7, 64.3) | 20.1 (19.0, 21.2) | 9.2 (8.5, 10.1) | 5.0 (4.4, 5.6) | 2.7 (2.3, 3.2) |
|  | n=1856 | n=3161 | n=1008 | n=464 | n=250 | n=134 |
